# Methadone and Suboxone^®^ mentions on Twitter: Thematic and Sentiment Analysis

**DOI:** 10.1101/2020.06.19.20135962

**Authors:** M Chenworth, J Perrone, JS Love, R Graves, W Hogg-Bremer, A Sarker

## Abstract

**Background:** Methadone and buprenorphine-naloxone (Suboxone®) have been discussed and compared extensively in the medical literature as effective treatments for opioid use disorder (OUD). While the evidence basis for the use of these medications is very favorable, less is known about the perceptions of these medications within the general public.

**Objective:** This study aimed to use social media, specifically Twitter, to assess the public perception of these medications, and to compare the discussion content between each medication based on theme, subtheme, and sentiment.

**Methods:** We conducted a mixed methods descriptive study analyzing individual microposts (“tweets”) that mentioned “*methadone*” or “*suboxone*”. We then categorized these tweets into themes and subthemes, as well as by sentiment and personal experience, and compared the information posted about these two medications, including in tweets that mentioned both medications.

**Results:** We analyzed 900 tweets, most of which related to *access* (13.8% for methadone; 12.9% for suboxone®), *stigma* (15.3%; 14.0%), and *OUD treatment* (11.5%; 5.4%). Only a small proportion of tweets (16.4*%* for suboxone® and 9.3% for methadone) expressed positive sentiments about the medications, with few tweets describing personal experiences. Tweets mentioning both medications primarily discussed MOUD in general, rather than comparing the two medications directly.

**Conclusions:** Twitter content about methadone and suboxone are similar, with the same major themes and similar sub-themes. Despite the proven effectiveness of these medications, there was little dialogue related to their benefits or efficacy in the treatment of opioid use disorder. Perceptions of these medications may contribute to their underutilization in combatting opioid use disorder.

## Introduction

Medications for opioid use disorder (MOUD), methadone, buprenorphine, and extended-release naltrexone, are all FDA-approved for the treatment of opioid use disorder (OUD) and have been shown to reduce all-cause mortality, overdose-related mortality and transmission of infectious diseases, while increasing treatment retention and improving long-term outcomes [1]. Despite their proven efficacy, these medications are underutilized for reasons such as treatment access barriers, stigma and misperceptions about their role in recovery. Little is known about the public perception or opinion about these medications and whether the experiences associated with these medications differ, although these factors may also be associated with their underutilization. Both methadone and buprenorphine are agonist therapies, while naltrexone is an opioid antagonist; due to these parallel modes of action, comparative effectiveness studies tend to focus on methadone and buprenorphine [2,3]. Methadone is a long-acting full agonist at the mu-receptor. In contrast, buprenorphine is a partial mu agonist which has a ceiling effect, thus, limiting euphoria and overdose. To mitigate the abuse liability, buprenorphine is combined with naloxone, and this combination medication is known by the trade name *Suboxone®*. There is, however, concern that buprenorphine can be diverted, and physicians are required to undergo an additional 8 hours of training in order to prescribe buprenorphine.

Past research to understand the perceptions about MOUDs have focused on interviewing individuals struggling with OUD, but such studies have narrow scopes, lack diverse cohorts and are expensive to conduct or coordinate [4]. Social media, due to its widespread use, offers the opportunity to obtain and study discussions and perceptions associated with specific topics, including health-related topics. One popular social media platform that has been used extensively in past research related to substance use is Twitter. Twitter is a *microblogging* website and has become a popular resource for individuals to access news, expand their social networks, and engage in discussions via publicly visible posts. Twitter has 326 million monthly active users, 100 million daily active users, and 500 million tweets per day [5]. Since Twitter microblogs are mostly publicly available, it has increasingly been used to investigate attitudes and perceptions, real-time content, and posts on specific topics through keywords and hashtags. Various health and public health-related topics have been studied in the past using Twitter, including topics associated with OUD and substance use disorder [6-9]. In this paper, we use data collected from Twitter to investigate the public perceptions of both buprenorphine-naloxone and methadone, and to compare the discussions of both medications by theme, sub-theme, and sentiment.

## Methods

### Data collection and preparation

We conducted a mixed-methods descriptive study with the aid of natural language processing (NLP) methods to analyze public tweets mentioning methadone and buprenorphine/naloxone. We used the Twitter application programming interface (API) to collect the data using the generic and brand names (Methadose® and Suboxone®) as keywords. Since medication names are often misspelled on Twitter, we used a spelling variant generator [10] to include misspellings of the medication names in the collection process. We found that in the case of methadone, the generic name was very commonly used in discussions, while trade names were rarely used. In the case of buprenorphine-naloxone, the trade name, Suboxone®, was most commonly used (Figure 1). Therefore, throughout the rest of this paper, we use the terms methadone and Suboxone® to refer to these medications. During the analyses, integer IDs were used for the tweets and the users in order to hide the user handles of the original posters from the annotators. We performed further filtering to clean the data, which involved removing duplicates, non-English tweets, very short tweets (*e*.*g*., those consisting of emojis only), and tweets that had too many unreadable symbols/emojis. Two levels of filtering were required for this—first, automatic filtering using NLP methods, and second, manual filtering during the annotation process. We only used publicly available posts for this study, and the study was reviewed and approved by the Institutional Review Board of the researchers’ home institution.

**Figure 1:**
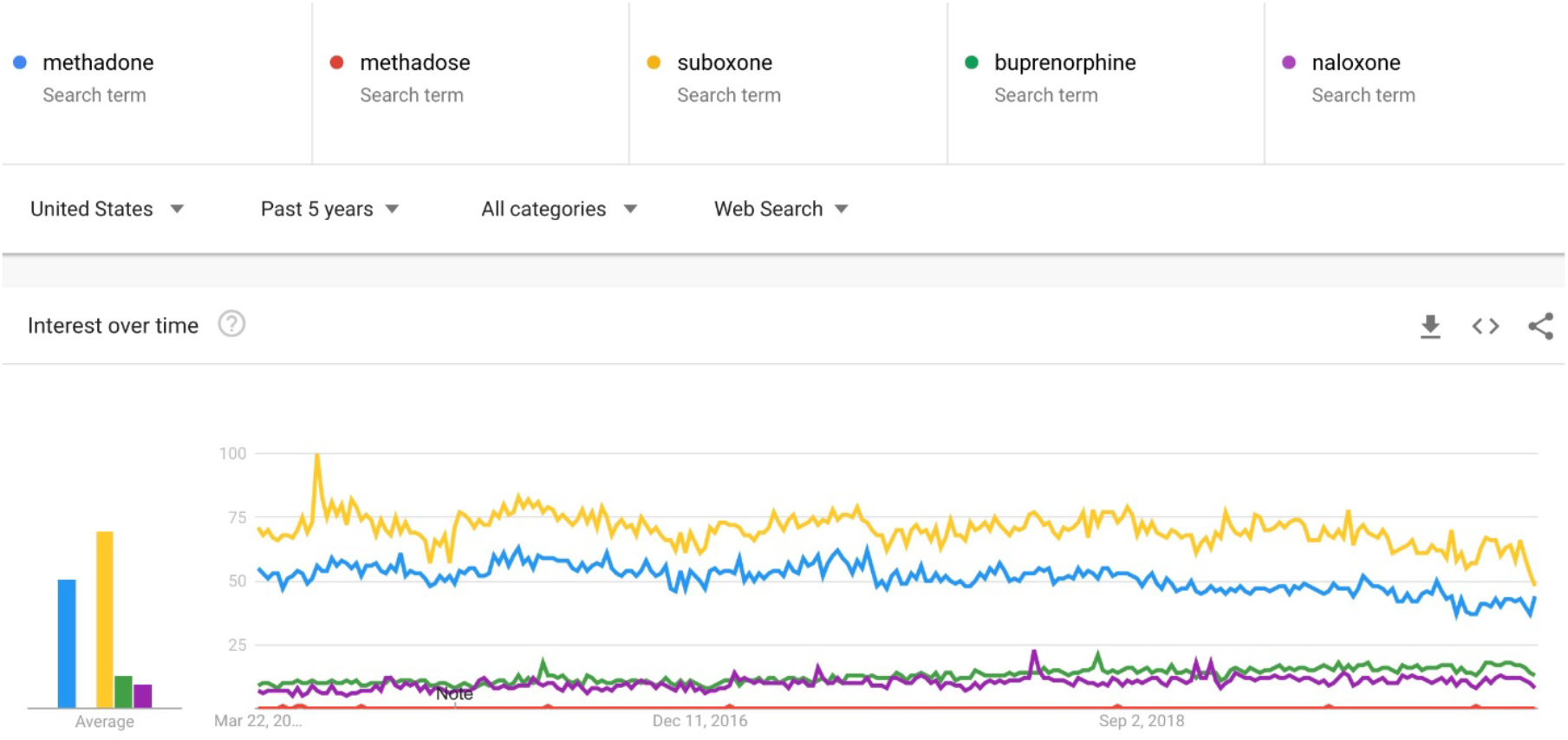
Google trends for the search terms ‘methadone’, ‘methadose’, ‘suboxone’, ‘buprenorphine’ and ‘naloxone over the last five years. The terms ‘methadone’ and ‘suboxone’ are most commonly used.

### Data annotation and analysis

From the collected and filtered Tweets, we drew a random sample for manual analysis. We used an adapted thematic analysis [11] to categorize the tweets. A sample of 100-150 tweets mentioning each medication were first reviewed to find common topics, and then these topics were coalesced first into broad *themes* that represented the high-level information contents of the tweets. Following this, we manually categorized the tweets within each theme into fine-grained *topics* or *sub-themes*. Each tweet could have one or more themes (including the *uncategorized* or *unrelated* themes) and zero or more sub-themes. The annotations were performed by three annotators (MC, AS and WH), with a small number (n=100) of overlapping tweets, which were used to compute inter-annotator agreements (IAA). We computed mean pair-wise IAA using Cohen’s kappa [12]. The disagreements between the annotators were resolved by AS.

We also coded the tweets based on the sentiment expressed in the context of the medication (negative, positive and neutral) and whether they represented personal experiences or not (n=900). Tweets considered positive referred to the MOUD in a positive light (*e*.*g*., the benefits of the MOUD), while tweets categorized as negative referred to the MOUD in a negative light (*e*.*g*., expressing stigma associated with the MOUD). Neutral tweets mentioned the MOUD but did not express positive or negative sentiments; they were often related to medical education, listing drugs, or were unrelated to MOUD discussion.

Following this manual categorization, we compared the themes, topics, and sentiments between the methadone and Suboxone® tweets (n=800). We computed the proportions of these themes across the annotated tweets and compared the differences in the proportions using a two-tailed test for proportions. We then performed descriptive analyses of the tweets belonging to each theme, particularly for the more frequently occurring ones, to better describe their contents in terms of sub-themes and expressed sentiments.

We qualitatively analyzed a sample of tweets that mentioned both Suboxone® and methadone to gain an understanding of *if* and *how* they are compared in those tweets. We categorized tweets as *pro-Suboxone®, pro-methadone, anti-Suboxone®, anti-methadone*, or *equal*. Tweets were categorized as pro-Suboxone® or pro-methadone if they mentioned the benefits of one medication compared to the other, as anti-Suboxone® or anti-methadone if the tweet mentioned downsides of one medication compared to the other, or equal if the tweet mentioned both medications in the same way. We further categorized these tweets in terms of whether the MOUDs were expressed in a positive way (pro-MOUD) or a negative way (anti-MOUD); tweets that were not obviously positive or negative were categorized as neutral.

## Results

### Manual categorization

We analyzed a total of 900 tweets, 450 for each medication. MC analyzed/annotated 400 tweets, derived the initial themes and sub-themes, and outlined the annotation guidelines. AS annotated 200 tweets (100 for each medication) and WHB annotated 500 tweets (250 for each medication), including 100 overlapping tweets with those annotated by MC. Average pair-wise IAA between the three annotators was κ = 0.767 (Cohen’s kappa[12]; pair-wise IAAs: 0.765,0.754, 0.790), which can be considered to be substantial agreement [13].

The methadone and Suboxone® tweet samples contained similar overall themes, though the more specific topics or sub-themes differed. In total, we included 11 themes. Table 1 provides definitions of the themes and samples of these tweets, while Table 2 provides the percentage of methadone and Suboxone® tweets belonging to these themes (Tables 1 and 2). The top three most frequently occurring themes, other than *uncategorized* or *unrelated*, were *access* (13.8% for methadone; 12.9% for suboxone®), *stigma* (15.3%; 14.0%), and *OUD treatment* (11.5%; 5.4%). Other themes included *pain management, tapering/withdrawal, safety/side effects, greed/corruption, continued drug use*, and *misuse/diversion*. Figure 2 visually depicts the distributions of the themes across the two medications.

**Table 1:**
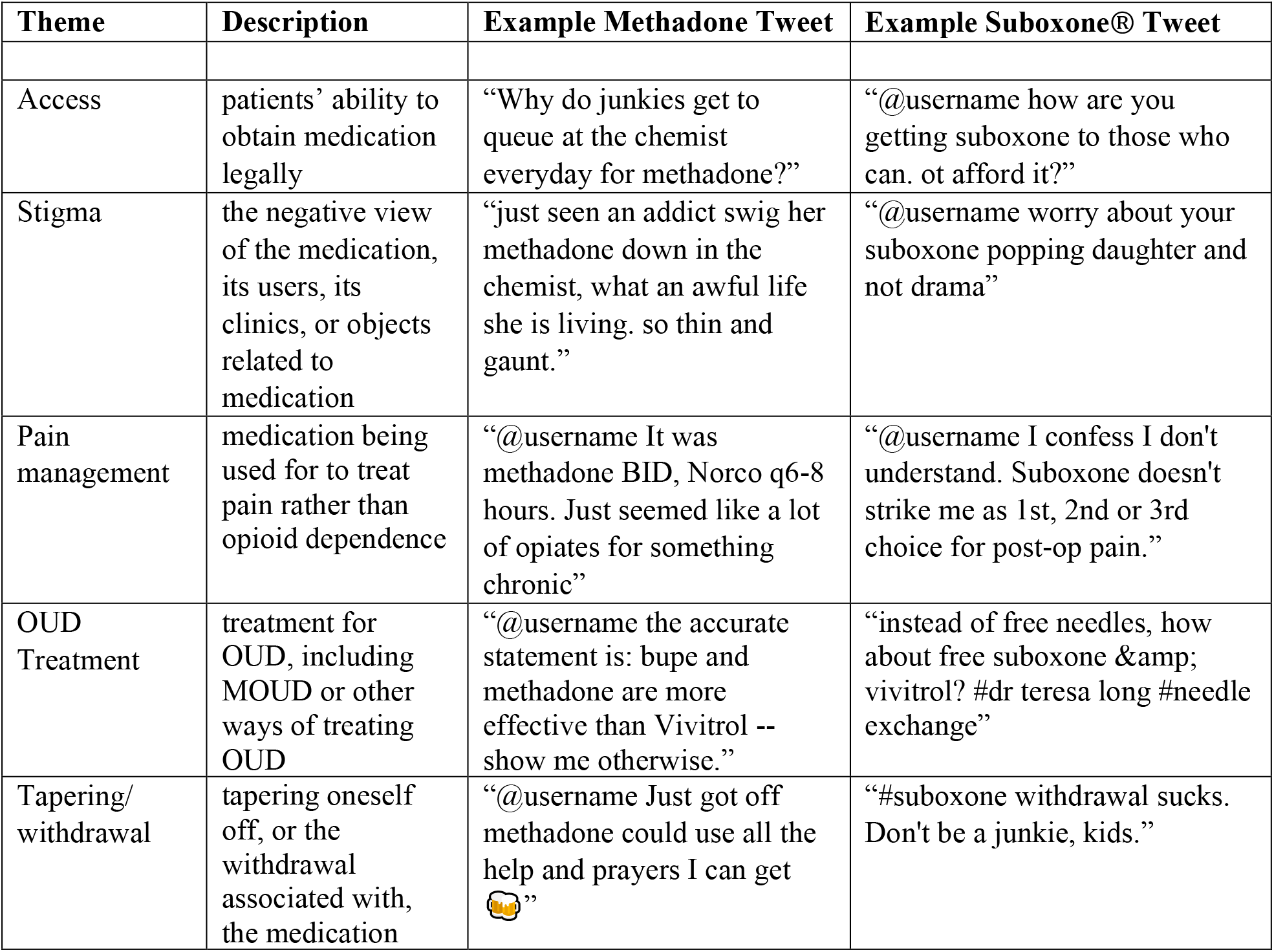
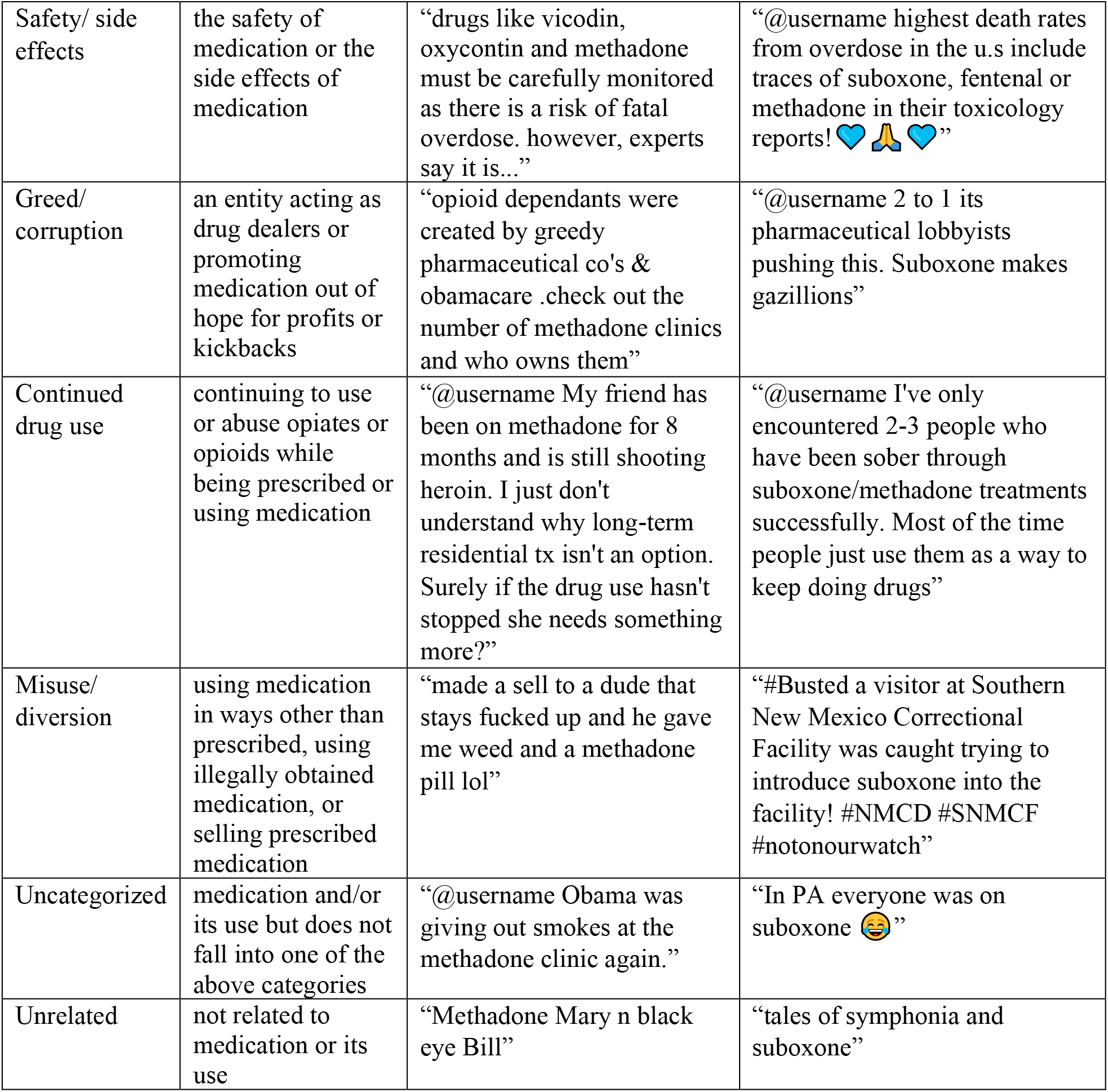
Description of themes associated with tweets mentioning suboxone® and methadone

**Table 2:** Analysis of themes associated with tweets mentioning suboxone® and methadone

| Theme | Methadone Tweets<br>(N=250) | Suboxone® Tweets<br>(N=250) | P |
| --- | --- | --- | --- |
|  | n (%) | n (%) |  |
| Access | 35 (13.8) | 32 (12.9) | .70 |
| Stigma | 35 (15.3) | 35 (14.0) | .70 |
| Pain management | 13 (5.0) | 16 (6.3) | .57 |
| OUD Treatment | 29 (11.5) | 38 (15.4) | .24 |
| Tapering/ withdrawal | 7 (2.7) | 15 (6.1) | .08 |
| Safety/ side effects | 13 (5.0) | 9 (3.6) | .38 |
| Greed/ corruption | 12 (4.7) | 31 (12.2) | .002<sup>a</sup> |
| Continued drug use | 4 (1.6) | 4 (1.6) | 1 |
| Misuse/ diversion | 5 (1.8) | 19 (7.7) | .003<sup>a</sup> |
| Uncategorized | 32 (12.6) | 44 (17.4) | .14 |
| Unrelated | 65 (26.0) | 7 (2.7) | <.001<sup>a</sup> |

<sup>a</sup>statistically significant difference

**Figure 2:**
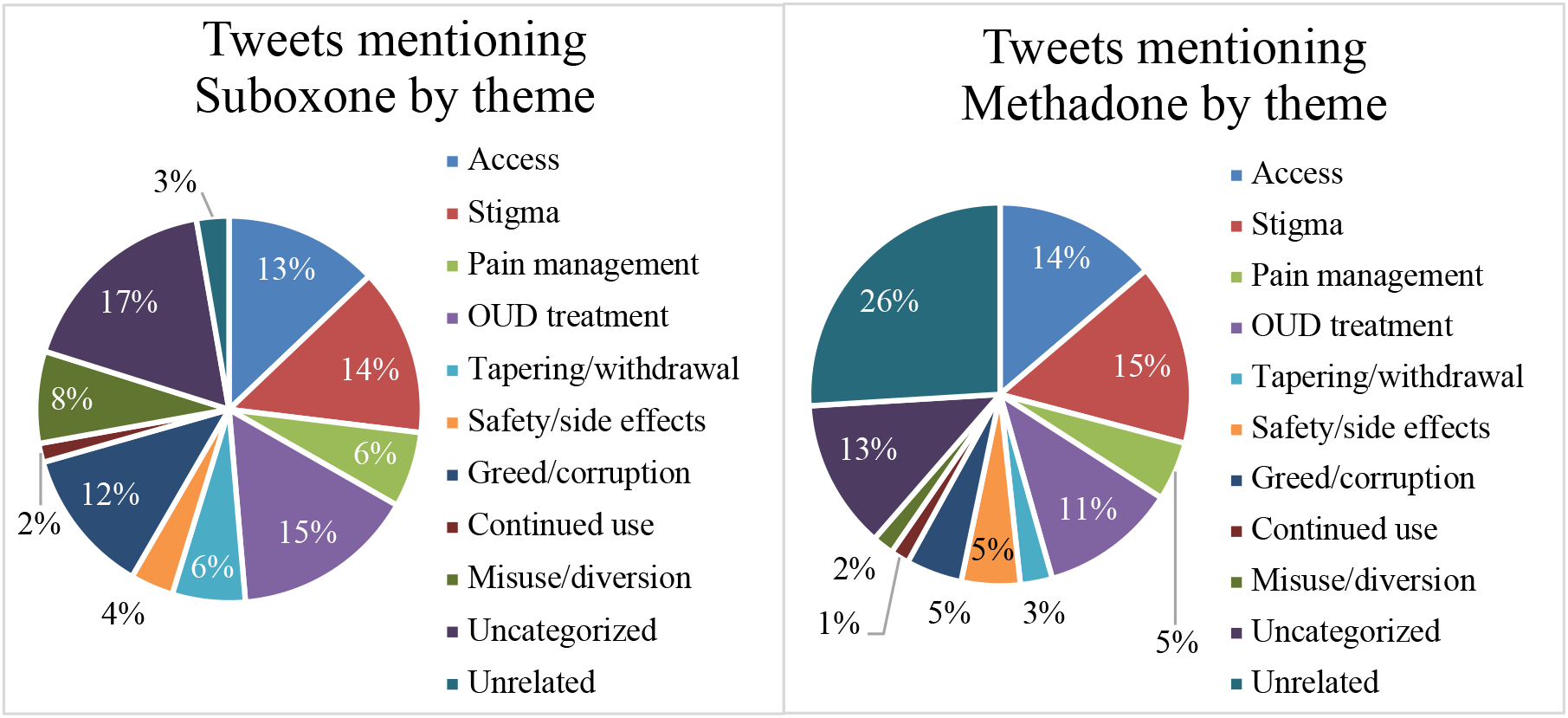
Distribution of themes in tweets mentioning Suboxone® and methadone.

Most tweets were not from the perspective of personal experience, with only 15.7% (20.4% for Suboxone® and 10.9% for methadone) of the analyzed tweets describing firsthand experiences and 6.3% (5.8% for Suboxone® and 6.9% for methadone) of tweets referring to the experiences of someone close to the poster, whether a family member, friend, or patient (n=900).

### Qualitative analysis

The tweets belonging to the *access* theme revealed some important issues that act as barriers to treatment for patients with OUD. These tweets typically presented the users’ views about obtaining the particular MOUD legally, difficulties faced when trying to access them, and often mentioned medication expense and cost coverage. Many tweets about Suboxone® within this theme focused on the high cost of this medication and the need for insurance, which act as barriers to access. Similarly, a portion of methadone mentions within this theme discussed whether the government covered the cost of the medication. Methadone discussions also often described the need for OUD patients to wait in line at designated clinics to receive methadone doses, including firsthand experiences posted by patients (Table 3). This need for queuing at clinics and wait while doses are verified and distributed is often described in medical literature as a barrier to treatment for patients, as well as a reason why zoning is difficult to obtain for methadone clinics [1]. Suboxone® tweets included those posted by physicians, who described the barriers faced by them, particularly the requirement to have a special license (the “*x waiver*”) to be able to prescribe the medication (Table 4). This x-waiver is extra training required by the government that prescribers must undergo before being allowed to prescribe buprenorphine, which ultimately limits patient access to this medication, as only about 5% of providers are x-waivered [14].

**Table 3:**
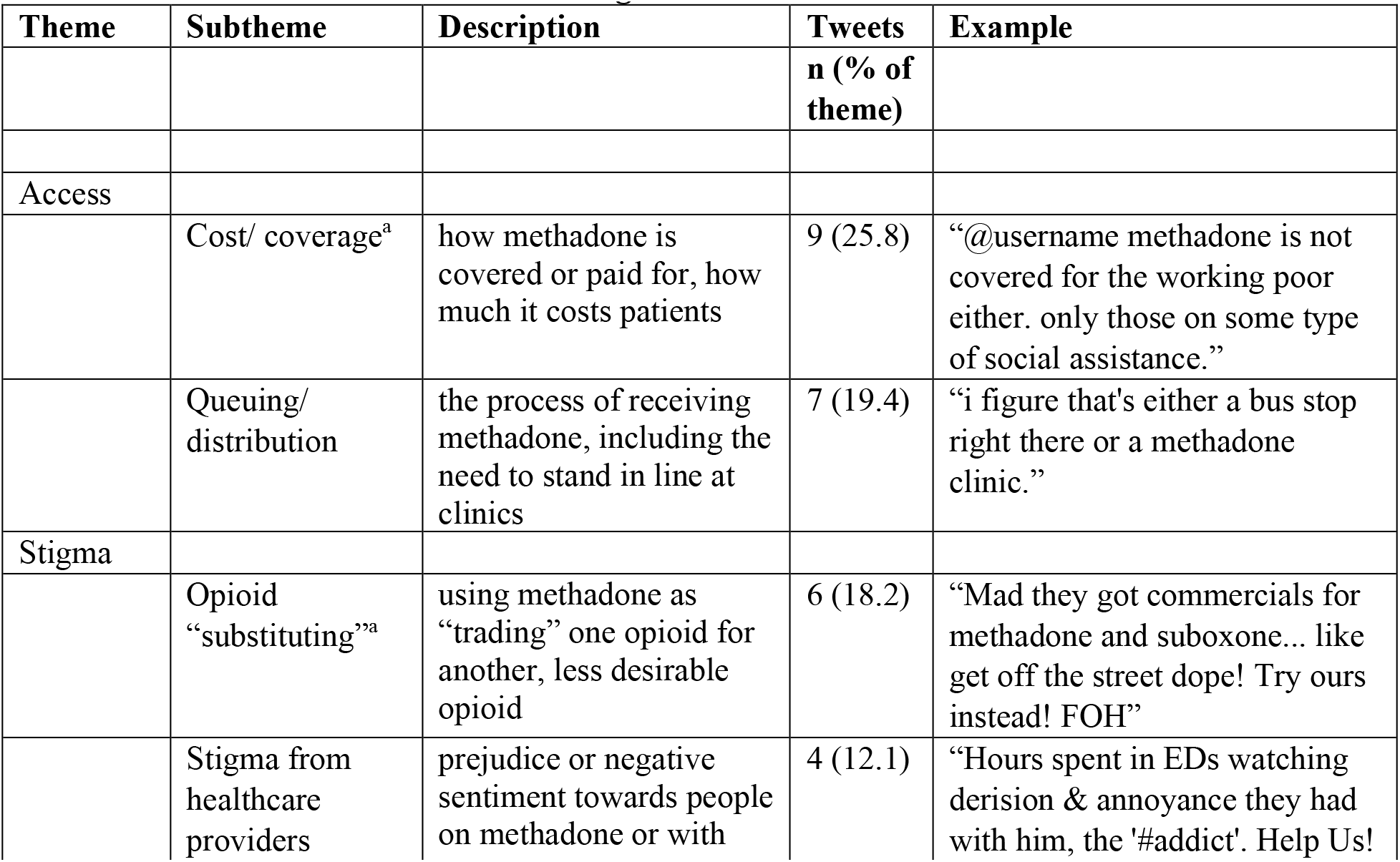
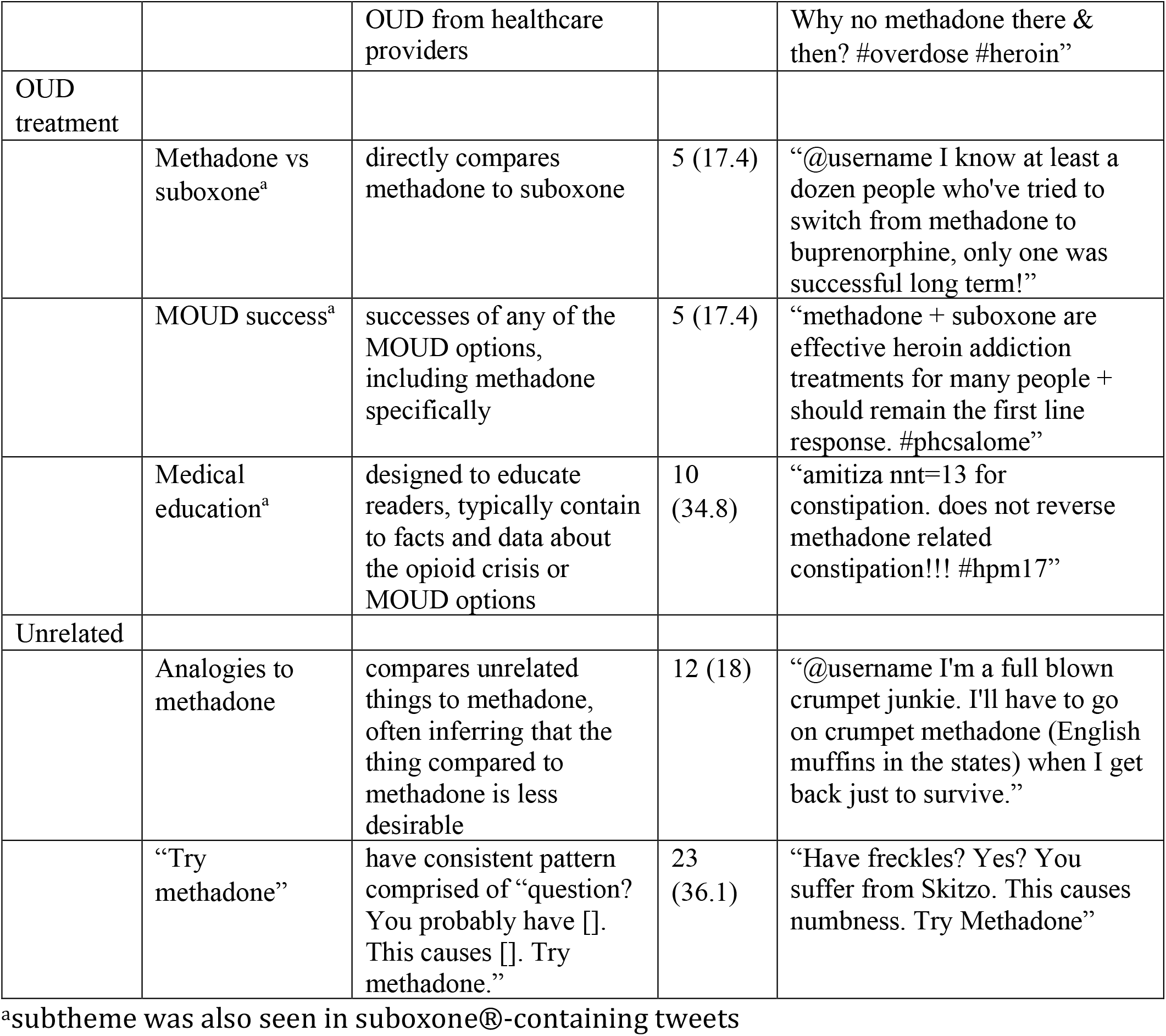
Subthemes of tweets mentioning methadone.

**Table 4:** Subthemes of tweets mentioning suboxone®.

| Theme | Subtheme | Description | Tweets n (% of theme) | Example |
| --- | --- | --- | --- | --- |
| Access |  |  |  |  |
|  | Cost/ Coverage<sup>a</sup> | how suboxone is covered or paid for, how much it costs patients | 13 (40.7) | “@username suboxone is very expensive, came to the u.s. from europe, initial training was data 2000 for an outpatient detox.” |
|  | MD awareness/ licensing | difficulties in obtaining suboxone due to physician awareness, prescribing, or licensing issues | 9 (29.6) | “@username but there are only a few docs who can give suboxone to people and they are restricted to 30” |
| Stigma |  |  |  |  |
|  | Opioid “substituting”<sup>a</sup> | using suboxone as “trading” one opioid for another, less desirable opioid | 10 (28.1) | “Mad they got commercials for methadone and suboxone... like get off the street dope! Try ours instead! FOH” |
|  | Myths | fallacies related to suboxone and the opioid epidemic | 9 (25) | “@username he was on suboxone which has links to aggressive and violent behavior.” |
|  | Suboxone “addiction” | suboxone treatment as “suboxone addiction” or refers to the addictive properties of suboxone | 5 (15.6) | “@username suboxone is one of the to drugs we detox patients from nowadays. #suboxoneaddiction” |
| OUD treatment |  |  |  |  |
|  | Methadone vs suboxone<sup>a</sup> | directly compares methadone to suboxone | 5 (12.8) | “@username Yes suboxone is a miracle drug to treat the crisis / methadone clinics should be shut down : have big lobby group” |
|  | MOUD success<sup>a</sup> | successes using any of the MOUD options, including specific successes from suboxone | 11 (28.2) | “@username i have 4 years 5 months and 13 days sober... suboxone saved my life for the better. amen to that! #beingthankful” |
|  | Medical education<sup>a</sup> | designed to educate readers, typically contain to facts and data about the opioid crisis or MOUD options | 9 (23.1) | “@username its already available some places. nalaxone blocks opiate receptors in brain. just like suboxone” |
| Tapering/ Withdrawal |  |  |  |  |
|  | Alternatives | alternatives to treat the opioid withdrawal (including kratom and cannabis) | 5 (35.7) | “will #kratom help ween of suboxone???” |

<sup>a</sup>subtheme was also seen in methadone-containing tweets

The *stigma* theme encompassed tweets that alluded to the public’s negative view about the particular medication. This included tweets that directly referenced instances of feeling stigmatized or judged based on the use of MOUD as well as tweets that indirectly referenced the stigma by referring to the medication, its users, or objects related to the use of the medication (such as Suboxone® wrappers) in a negative light. Both methadone and Suboxone® tweets asserted the idea that using methadone or Suboxone® is simply “substituting one drug or addiction for another” and that abstinence from all opioids is the only acceptable goal for people with OUD. We categorized such tweets as “*opioid substituting*” (Table 3, Table 4). Methadone tweets also described situations where individuals experienced stigma from healthcare providers, while Suboxone® tweets mentioned stigma associated with myths (such as that Suboxone® makes a person more violent, prescription opioids cannot cause addiction, or anti-psychotic drugs are included in Suboxone®), referenced the idea of a “*suboxone addiction*”, or stated the addictive properties of Suboxone®.

The *OUD treatment* theme encompassed tweets that discussed treatment-related chatter for OUD, including MOUD and other ways of treating OUD. Many compared methadone and Suboxone® directly, offered opinions on which is better, shared stories of success related to these medications, or denounced these options altogether (Table 3, Table 4). Many of these tweets also focused on medical education or awareness, designed to educate and typically referring to facts or data about the opioid crisis or MOUD options but not imbued with personal opinion or experience.

### Sentiment analysis

Most methadone tweets were categorized as neutral (49.1%; n = 450), while the majority of Suboxone® tweets were categorized as negative (55.8%; n = 450) (Figure 3). While the “positive” categorization made up the smallest proportion of tweets in both groups, the Suboxone® sample had more tweets categorized as positive than the methadone sample (16.4*% vs*. 9.3%). In both tweet samples, “positive” tweets discussed the medications’ benefits, success stories, or expressed the idea that the MOUD is good for society, while “negative” tweets often mentioned side effects, toxicity, stories of failure or continued abuse, or experience of stigma. Neutral tweets were often related to medical education, listing drugs, or were unrelated to MOUD discussion.

**Figure 3:**
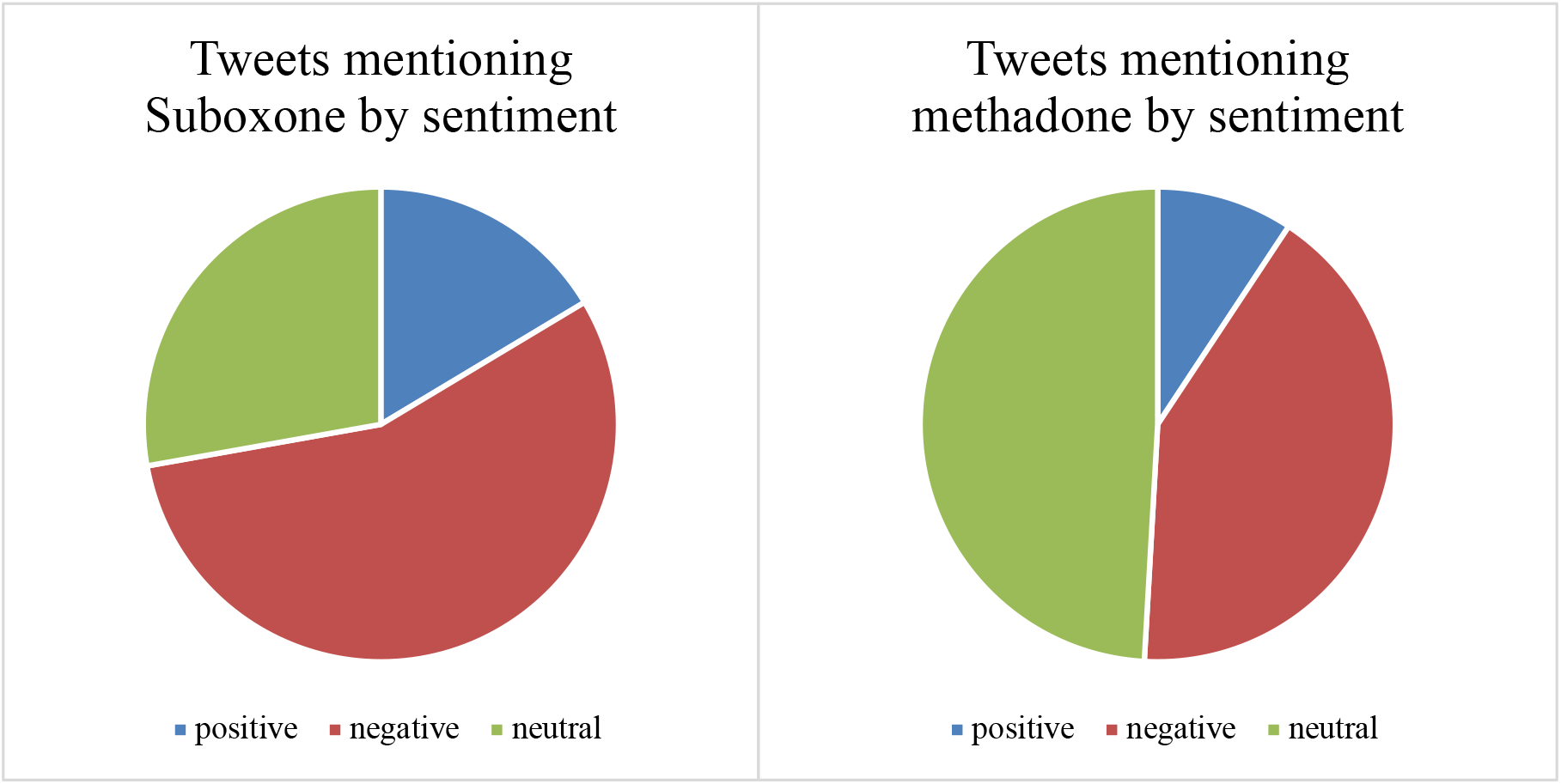
Distribution of sentiments in tweets mentioning Suboxone® and methadone.

### Analysis of tweets mentioning both medications

In tweets that mentioned both Suboxone® and methadone, 84.0% (168/200) mentioned the medications in the same light, thus being categorized as *equal* (Figure 4). Of these 168 tweets, 39 tweets were *anti-MOUD* (23.0%), 64 were *neutral* (38.0%), and 65 were *pro-MOUD* (39.0%). The remainder of the tweets were more likely to mention the benefits of one medication over the other, with 15 tweets being *pro-Suboxone®* (8%) and 7 tweets being *pro-methadone* (4%). There were only a small number of tweets that were pointedly against one medication (6 *anti-Suboxone®* tweets [3%]; 4 *anti-methadone* tweets [2%]).

**Figure 4:**
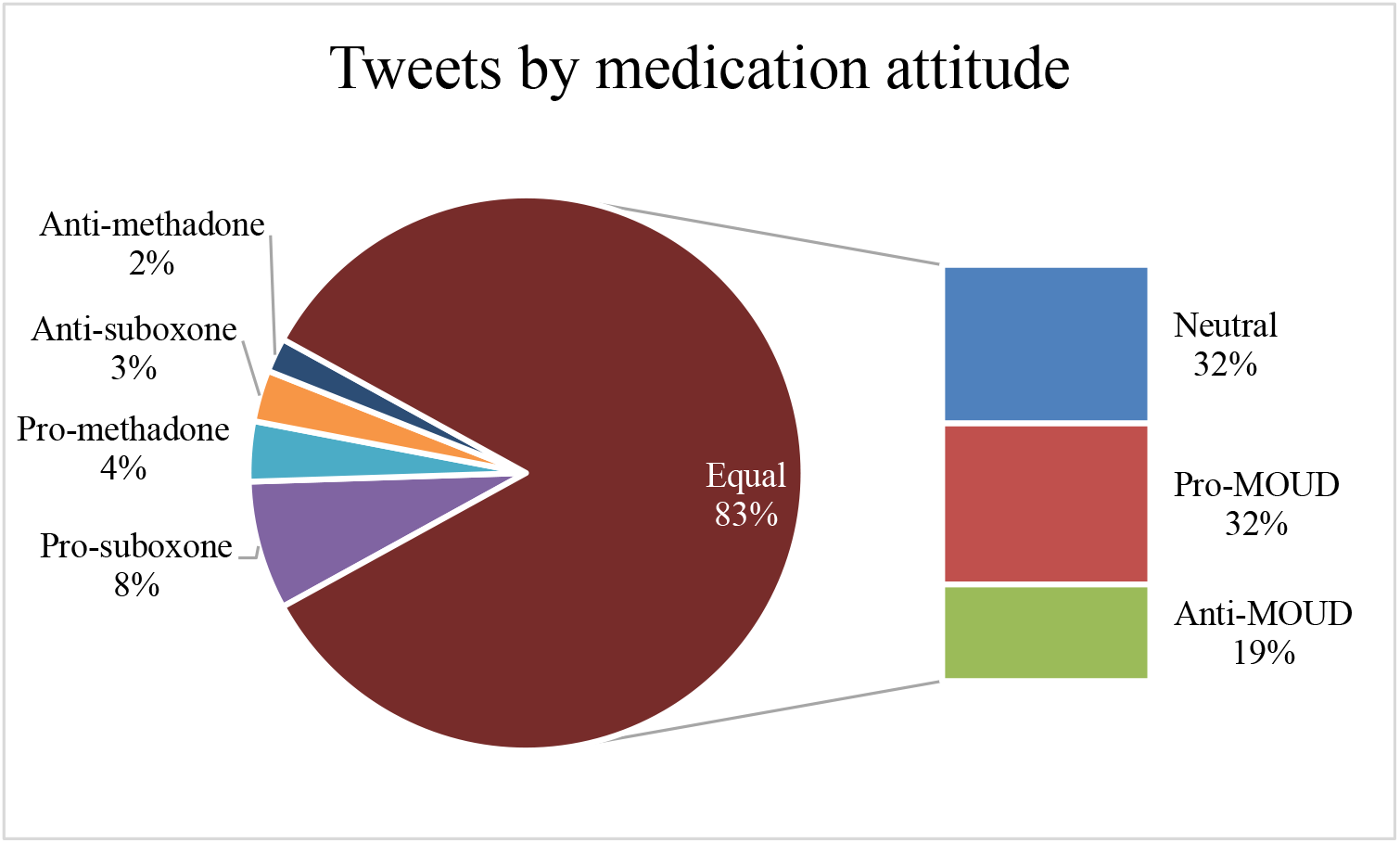
Tweets mentioning both “methadone” and “suboxone” by attitude on medications.

## Discussion

### Benefits of and barriers to MOUD treatment

Much of the medical literature has focused on the perception of benefits and barriers as they relate to MOUD-based treatment from the viewpoints of doctors and the healthcare systems rather than from the perspectives of the public. The report from the Committee on Medication Assisted Treatment for OUD in 2019 identified the major barriers to MOUD as being misunderstanding and stigma related to OUD and its treatment, inadequate education of professionals meant to treat these patients, current regulations surrounding methadone and buprenorphine, and the fragmented system of care for OUD patients [1]. This summary document also highlighted the effectiveness of these medications in reducing all-cause mortality, overdose-related mortality, and transmission of infectious diseases, as well as increasing treatment retention, and improving long-term outcomes in OUD patients [1]. While many of the methadone- and Suboxone®-related themes we discovered were similar to those discussed in the medical literature, the discussions on Twitter did not include as many of the benefits that these medications have in treating OUD. The sub-theme “MOUD success” only comprised 2% of the methadone tweets and 5.5% of the Suboxone® tweets, suggesting a relative lack of public awareness to the benefits of both methadone and Suboxone®. As an increasing number of individuals search for relevant health-related information on social media, this relative lack of discussion of the benefits of methadone and Suboxone® on this platform may skew public opinion of these medications, and hinder efforts to engage patients in evidence-based treatments to address the opioid crisis. This also highlights one area of potential intervention by the healthcare community: there is a role for providers to engage with the public and participate in discussions surrounding MOUD on social media to share success stories and data regarding the benefits of these medications in treating OUD. There is some evidence of this in our samples, with 2% (8) of the 400 tweets analyzed mentioning a doctor-patient or provider-client relationship with individuals with OUD. Interestingly, tweets that mentioned both medications were significantly more likely to be positive than tweets that only mentioned one medication (*P*<0.0001), suggesting that users that are aware of the benefits of these medications discuss these medications together.

One important observation of the discussion surrounding barriers to treatment with these medications was the *stigma* theme. One sub-theme that arose in both the Suboxone® and methadone tweets was “*opioid substitution*”—the perception that these medications lead patients with OUD to exchange one opioid addiction for another more socially acceptable one. This is a barrier that makes addressing the opioid crisis more difficult, as discussions online may be the first exposure for a person with OUD when considering treatment. In fact, information seeking posts were found in both our Suboxone® and methadone tweet samples, indicating that some people use these public discussions to obtain information on MOUD and to obtain community information about these medications. Similarly, another concept from the *stigma* theme was tweets mentioning stigma from healthcare providers, including accounts of patients being told to leave the emergency department (ED) to go to the methadone clinic, as the ED does not prescribe methadone. This is unfortunately a barrier known in OUD treatment, as some healthcare providers have demonstrated negative attitudes towards patients with OUD [15].

Experiencing this stigma as well as reading about it online could prevent patients with OUD from seeking treatment with MOUD for fear of poor treatment from the healthcare encounter. In fact, this concept was present in the discussions of both Suboxone® and methadone samples.

### Using social media to assess public perceptions

Past research on analyzing public perceptions and discussions surrounding methadone and Suboxone® focused primarily on conducting interviews with people struggling with OUD in inpatient treatment facilities. For example, in 2016 Yarborough and colleagues grouped the discussions they had with adults with opioid dependence (n = 283) into themes they called “*areas of consideration*” for MOUD decision making. These included many of the same themes we found in our Twitter discussion including awareness of treatment options, stigma associated with methadone clinics, fear of continued addiction and perceived difficulty of withdrawal, and pain control [16]. Other “areas of consideration” included expectations and goals for duration of treatment and abstinence, prior experience with buprenorphine or methadone, and the need for accountability and structured support. While our analysis included only 1,100 tweets mentioning Suboxone®, methadone, or both, it is apparent that social media in general, and Twitter specifically, can be utilized to analyze far more individuals’ perceptions on MOUDs than traditional studies involving individual interviews.

The studies based on face-to-face interviews were also limited in scope by only interviewing people currently in OUD treatment; thus, excluding the perceptions of the general population, and the families and friends of patients treated for OUD. A study conducted by Brown and Altice in 2014, investigated the possibility of garnering information regarding buprenorphine/naloxone from online discussion boards, which were public and included discussions posted by people with OUD and the general public. The authors categorized the discussion into themes that facilitated self-management with buprenorphine/naloxone. Although the study excluded methadone, the themes discovered by the authors were similar to those found in our current study, including the need for a ready supply of the medication from a variety of sources, distrust of buprenorphine prescribers and the pharmaceutical companies, and a desire to become completely “substance free” [17].

### Limitations and Future Directions

Due to the use of social media data only, our analyses were limited to individuals who were on Twitter and who actively self-reported information. This limits the generalizability of our findings and is the major limitation of our study. Twitter users tend to be younger and more racially diverse than the general population. Despite this limitation, Twitter has previously been used to examine attitudes and perceptions of other topics and those attitudes have been shown to be consistent with attitudes found using survey measures, and such a limitation also affects any survey-based studies [18]. For example, in 2018, Farhadloo and colleagues examined the Twitter discussion on Zika and compared that discussion to survey measures of Zika-related attitudes, knowledge, and behaviors and found associations between the two. They ultimately concluded that Twitter data could be used as a complementary source of information to gauge patterns of attitudes, knowledge, and behaviors in a population [19].

Another limitation of this study is its retrospective nature. Discussion on social media are constantly shifting and changing, and analysis of a sample of tweets can only offer a snapshot in time of the discussion. However, our samples were from tweets posted over a 9-year period and randomized for analysis, allowing our sample to represent nearly a decade of discussion. While the Twitter discussion and thus public perception of these medications may have shifted in the time it took to analyze the tweets, it is unlikely that the discussion would be significantly different from the past 9 years.

Our investigation also did not include demographic data of the users, geographical distributions of the posts, or information regarding who posted the tweets. It is possible that our samples of tweets came from specific geographical areas of the country due to higher levels of discussion of these topics in those areas, or that participants in the discussion had similar backgrounds that we did not see or analyze.

As described earlier, one future direction of this work would be to use social media like Twitter to engage with the public discussion of methadone and Suboxone®. This will allow practitioners to highlight aspects of the discussion that have not been covered, such as the numerous benefits of MOUD and the strong data behind it, as well as address some of the myths and stigma surrounding these medications. In the future, it will be interesting to evaluate whether interventions by physicians can change the public discussion and perception about these medications, and if localized interventions can change prescription practices. Compared to all the methadone and Suboxone® related information available on Twitter, we only studied a small sample. Manual characterization of Twitter data, although very accurate and fine-grained, is time-consuming. In the future, building on our extensive past work on social media mining for health, we will conduct automated analyses of much larger volumes of social media data about these medications.

## Conclusions

Our analyses of a sample of Twitter posts mentioning methadone and Suboxone® demonstrates that the discussions regarding the two medications are very similar, with the same major themes and many of the same sub-themes. Additionally, our analyses showed that some of the discussions on Twitter parallel that of the medical community, such as the barriers to implementing and using MOUDs for treatment. However, the Twitter discussions contained little discourse related to the benefits of these medications. This suggests a relative paucity of public awareness of benefits of methadone and Suboxone® and offers an opportunity for public education in the future. Although these discussions are happening spontaneously on social media, it may be possible for the medical community to actively contribute to these discussions and raise awareness of the scientific evidence of the benefit of these medications and disseminate stories of success.

## Data Availability

This data set will not be publicly shared in order to protect the identities of the original posters.

## Acknowledgements

Research reported in this publication was supported by NIDA of the NIH under award number R01DA046619. The content is solely the responsibility of the authors and does not necessarily represent the official views of the NIH.

## Abbreviations

API: application programming interface
ED: emergency department
IAA: inter-annotator agreements
MOUD: medications for opioid use disorder
OUD: opioid use disorder

